# Multi-Sequence Average Templates for Aging and Neurodegenerative Disease Populations

**DOI:** 10.1101/2021.06.28.21259503

**Authors:** Mahsa Dadar, Richard Camicioli, Simon Duchesne, For the CCNA Group

## Abstract

Magnetic resonance image (MRI) processing pipelines use average templates to enable standardization of individual MRIs in a common space. MNI-ICBM152 is currently used as the standard template by most MRI processing tools. However, MNI-ICBM152 represents an average of 152 healthy young adult brains and is vastly different from brains of patients with neurodegenerative diseases. In those populations, extensive atrophy might cause inevitable registration errors when using an average template of young healthy individuals for standardization. Disease-specific templates that represent the anatomical characteristics of the populations can reduce such errors and improve downstream driven estimates.

We present multi-sequence average templates for Alzheimer’s Dementia (AD), Fronto-temporal Dementia (FTD), Lewy Body Dementia (LBD), Mild Cognitive Impairment (MCI), cognitively intact and impaired Parkinson’s Disease patients (PD-CIE and PD-CI, respectively), individuals with Subjective Cognitive Impairment (SCI), AD with vascular contribution (V-AD), Vascular Mild Cognitive Impairment (V-MCI), Cognitively Intact Elderly (CIE) individuals, and a human phantom. We also provide separate templates for males and females to allow better representation of the diseases in each sex group.

## Background and Summary

Magnetic resonance imaging (MRI) brain templates (i.e. averages of multi-individual images, co-registered in a similar reference space) are widely used in image processing, for example as targets in registration and intensity normalization, as a common standard space enabling individual and population based comparisons in deformation/tensor or voxel based morphometry, and as the basis for segmentation techniques that rely on nonlinear registration ^1–4^. An example is the MNI-ICBM152, an average based on images from 152 healthy young adults, and one of the most popular templates in current use given its distribution in processing pipelines such as MINC, FSL, and SPM ^1–3^ that have been shared more than 45,000 times worldwide (Data from NITRC.org).

A common feature of existing averages such as the MNI-ICBM152 is their reliance on healthy, young brains, in addition to aggregating both sexes in the template generation process. However, in aging and populations with neurodegenerative diseases, ventricle enlargement, extensive levels of cortical and subcortical atrophy, as well as white matter hyperintensities (WMHs) create large degrees of difference between an individual’s MRI and such templates. We have shown in prior work that such differences significantly increase registration errors in some of these well-known image processing tools (e.g. ANTs, Elastix, FSL, MINC, and SPM) ^5^. These errors would inevitably propagate downstream to other pipeline steps, making the derived estimates inaccurate, if not unusable. Using population appropriate templates is therefore necessary to reduce such errors by improving the resulting registrations as well as all subsequent pipeline estimates.

Based on data from the Canadian Consortium for Neurodegeneration and Aging (CCNA) ^6^, a flagship study of the Canadian Institutes of Health Research, we present average templates for T1-weighted (T1w), T2-weighted (T2w), T2*-weighted, Proton Density (PD), and FLuid Attenuated Inversion Recovery (FLAIR) sequences in eleven diagnostic groups, including Alzheimer’s Dementia (AD), Fronto-temporal Dementia (FTD), Lewy Body Dementia (LBD), Mild Cognitive Impairment (MCI), cognitively intact and impaired Parkinson’s Disease patients (PD-CIE and PD-CI, respectively), individuals with Subjective Cognitive Impairment (SCI), Vascular Alzheimer’s Dementia (V-AD), Vascular Mild Cognitive Impairment (V-MCI), as well as Cognitively Intact Elderly (CIE) individuals and one human phantom ^7^. In addition to improving image processing outcomes, these templates capture anatomical characteristics for each disease cohort at the regional level. With multiple contrasts available providing different types of information, the various templates can be used to assess different aspects in each disease: i) T1w templates are useful for assessing fine anatomical details and estimating regional and global atrophy levels; ii) T2w/PD sequences are useful for skull segmentation, and assessment of deep gray matter structures, iii) FLAIR images can be used to detect WMHs and infarcts; and iv) T2* images can be used to identify microbleeds as well as hemorrhages.

There are significant sex and gender related differences in the prevalence, clinical outcomes, and response to treatments for these distinct neurodegenerative diseases (e.g. higher prevalence of Alzheimer’s disease in females and higher prevalence of Parkinson’s disease in males) ^8–10^. Sex-specific average templates would therefore be useful tools to represent and assess potential anatomical differences in patterns of atrophy in males and females. Thus, in addition to the disease-specific average templates combining male and female participants, we provide separate templates for males and females in each diagnostic category.

## Methods

### Data

We used data from the Comprehensive Assessment of Neurodegeneration and Dementia (COMPASS-ND) cohort of the CCNA, a national initiative to catalyze research on dementia ^6^. COMPASS-ND includes deeply phenotyped subjects with various forms of dementia and mild memory loss or concerns, along with cognitively intact elderly subjects. Ethical agreements were obtained at all respective sites. Written informed consent was obtained from all participants.

Clinical diagnoses were determined by participating clinicians based on longitudinal clinical, screening, and MRI findings (i.e. diagnosis reappraisal was performed using information from recruitment assessment, screening visit, clinical visit with physician input, and MRI). The diagnostic groups included, AD, CIE, FTD, LBD, MCI, PD-CIE, PD-MCI, PD-Dementia (for this study, PD-MCI and PD-Dementia groups were merged into one PD-CI group), SCI, V-AD, and V-MCI. For details on clinical group ascertainment, see Pieruccini-Faria et al. ^11^. A single cognitively healthy volunteer was also scanned as a human phantom multiple times across different centers for quality assurance purposes (more information on the SIMON human phantom dataset can be found in Duchesne et al. 2019 ^7^).

Table 1 summarizes the demographic characteristics of the participants used to generate each template.

**Table 1.** Demographic characteristics of the participants used to create the average templates.

| Measure | N |  |  | Age |  |  | P Value |
| --- | --- | --- | --- | --- | --- | --- | --- |
| Diagnosis | Total | Female | Male | Total | Female | Male |  |
| AD | 73 | 29 | 44 | 74.34 ± 7.58 | 73.09 ± 7.57 | 75.17 ± 7.55 | 0.25 |
| CIE | 94 | 76 | 18 | 70.18 ± 6.05 | 70.21 ± 6.03 | 70.03 ± 6.33 | 0.91 |
| FTD | 28 | 16 | 12 | 66.91 ± 8.29 | 65.95 ± 6.77 | 68.20 ± 10.15 | 0.49 |
| LBD | 21 | 2 | 19 | 72.25 ± 8.11 | 73.68 ± 2.52 | 72.10 ± 8.51 | 0.80 |
| MCI | 210 | 92 | 118 | 72.04 ± 6.66 | 71.43 ± 6.66 | 72.51 ± 6.65 | 0.24 |
| Mixed | 41 | 22 | 19 | 78.89 ± 6.63 | 80.45 ± 6.69 | 77.26 ± 6.32 | 0.12 |
| PD-CIE | 65 | 31 | 34 | 66.66 ± 6.91 | 67.79 ± 6.38 | 65.69 ± 7.28 | 0.22 |
| PD-CI | 45 | 7 | 38 | 72.01 ± 7.58 | 67.84 ± 13.01 | 72.75 ± 6.13 | 0.12 |
| SCI | 125 | 93 | 32 | 70.57 ± 5.91 | 70.92 ± 5.95 | 69.58 ± 5.75 | 0.27 |
| V-AD | 27 | 11 | 16 | 77.34 ± 7.07 | 76.47 ± 6.65 | 78.05 ± 7.53 | 0.56 |
| V-MCI | 135 | 61 | 74 | 76.22 ± 6.32 | 74.32 ± 6.21 | 77.78 ± 6.01 | 0.001 |
| SIMON | 68 | - | 68 | 44.75 ± 1.47 | - | 44.75 ± 1.47 | - |

All participants were scanned using the Canadian Dementia Imaging Protocol, a harmonized MRI protocol designed to reduce inter-scanner variability in multi-centric studies and which included the following sequences ^12^:

– 3D isotropic T1w scans (voxel size = 1.0 × 1.0 × 1.0 mm^3^) with an acceleration factor of 2 (Siemens: MP-RAGE-PAT: 2; GE: IR-FSPGR-ASSET 1.5; Philips: TFE-Sense: 2)
– Interleaved proton density/T2-weighted (PD/T2w) images (voxel size = 0.9 × 0.9 × 3 mm^3^), fat saturation, and an acceleration factor of 2.
– Fluid attenuated inversion recovery (T2w-FLAIR) images (voxel size = 0.9 × 0.9 × 3 mm^3^), fat saturation, and an acceleration factor of 2.
– T2* gradient echo images (voxel size = 0.9 × 0.9 × 3 mm^3^) and acceleration factor of 2.

Table 2 shows the acquisition parameters for each sequence and scanner manufacturer. A detailed description, exam cards, and operators’ manual are publicly available at: www.cdip-pcid.ca.

**Table 2.** Acquisition parameters of the CDIP protocol.

| Sequence | Scanner Model | Matrix | Resolution (mm<sup>3</sup>) | Number of Slices | TR (msec) | TE (msec) | TI (msec) | Flip Angle |
| --- | --- | --- | --- | --- | --- | --- | --- | --- |
| T1w | GE | 256 × 256 | 1.0 × 1.0 × 1.0 | 180 | 6.7 | 2.9 | 400 | 11 |
|  | Philips | 256 × 248 | 1.0 × 1.0 × 1.0 | 180 | 7.3 | 3.3 | 935 | 9 |
|  | Siemens | 256 × 256 | 1.0 × 1.0 × 1.0 | 192 | 2300 | 2.98 | - | 9 |
| T2w/PD | GE | 256 × 256 | 0.94 × 0.94 × 3.0 | 48 | 3000 | 11/85 | - | 125 |
|  | Philips | 256 × 254 | 0.94 × 0.94 × 3.0 | 48 | 3000 | 13/100 | - | 90 |
|  | Siemens | 256 × 256 | 0.94 × 0.94 × 3.0 | 48 | 3000 | 10/91 | - | 165 |
| FLAIR | GE | 256 × 256 | 0.94 × 0.94 × 3.0 | 48 | 9000 | 140 | 2500 | 125 |
|  | Philips | 256 × 224 | 0.94 × 0.94 × 3.0 | 48 | 9000 | 125 | 2500 | 150 |
|  | Siemens | 256 × 256 | 0.94 × 0.94 × 3.0 | 48 | 9000 | 123 | 2500 | 165 |
| T2* | GE | 256 × 256 | 0.94 × 0.94 × 3.0 | 48 | 650 | 20 | - | 20 |
|  | Philips | 256 × 256 | 0.94 × 0.94 × 3.0 | 48 | 650 | 20 | - | 20 |
|  | Siemens | 256 × 256 | 0.94 × 0.94 × 3.0 | 48 | 650 | 20 | - | 20 |

TR: repetition time; TE: echo time; TI: inversion time.

### Preprocessing

All images were pre-processed with image denoising ^13^, intensity non-uniformity correction ^14^, and image intensity normalization into a 0-100 range. The pre-processed images were then linearly ^5^ registered to the pseudo-Talairach space defined by the MNI-ICBM152-2009c template using a 9-parameter registration (three translation, three rotation, and three scaling parameters) ^15^. T2w, PD, FLAIR, and T2* images were also co-registered (rigid registration, 6 parameters) to the T1w images with a mutual information cost function.

### Template Generation

The method by Fonov et al. was used to generate unbiased templates for each diagnostic group for all participants, as well as each group but separately for males and females ^16,17^ (all except the LBD group in which there were only two female participants). This method has previously been used to generate templates in various studies, including the latest higher resolution version of the MNI-ICBM2009c template (http://nist.mni.mcgill.ca/?p=904) ^15,18^. In short, the pipeline implements a hierarchical nonlinear registration procedure using Automatic Nonlinear Image Matching and Anatomical Labelling (ANIMAL) ^19^, iteratively refining the previous registrations by reducing the step size (20 iterations in total, four iterations at each of the levels of 32, 16, 8, 4, and 2 mm, respectively) until convergence is reached. This process of increasingly refined iterative nonlinear registrations leads to average brains that reflect the anatomical characteristics of the population of interest with higher levels of anatomical detail ^17^. The higher resolution T1w images (isotropic 1mm^3^) were used to obtain the nonlinear transformations for creating the average templates. T2w, PD, FLAIR, and T2* templates were then created by combining their rigid to-T1w co-registration transformations with the nonlinear transformations based on the T1w images. All final templates were generated at 1mm^3^ isotropic resolution.

### FreeSurfer Segmentation

To appreciate differences between templates, we processed all T1w averages using *FreeSurfer* version 6.0.0 (*recon-all -all*). *FreeSurfer* provides a full processing stream for structural T1w data (https://surfer.nmr.mgh.harvard.edu/) ^20^. The final segmentation output (aseg.mgz) was then used to obtain volumetric information for each template based on the FreeSurfer look up table available at https://surfer.nmr.mgh.harvard.edu/fswiki/FsTutorial/AnatomicalROI/FreeSurferColorLUT.

### Data Records

The average template files for all groups and sequences are available in both compressed MINC ^21,22^ and NIfTI formats at G-Node (https://gin.g-node.org/mahsadadar/CDIP_Templates)^23^ as well as Zenodo (https://zenodo.org/record/5018356#.YNPNkExE3b0)^24^.

### Technical Validation

#### Quality Control

The quality of the registrations, pre-processed images, as well as the volumetric segmentations performed by FreeSurfer was visually assessed by an experience rater. All images passed this quality control step. Note that the provided data was already quality controlled by the CCNA imaging platform for presence of imaging artifacts, and only scans that had passed this quality control step were acquired and used for this study.

#### Templates

Figures 1 to 5 show axial slices of the T1w, T2w, T2star, PD, and FLAIR average templates for all 11 diagnostic groups, covering the brain at different levels. For more detailed figures of each template, see the supplementary materials (Figures S.1-S.11). As expected, CIE, PD-CIE, and MCI groups had smaller ventricles, with lower levels of atrophy compared with the cognitively impaired and dementia groups (Figure 1). FLAIR images of the vascular cohorts (i.e. Mixed, V-MCI, and V-AD) showed extensive levels of periventricular hyperintensities compared to other groups (Figure 2), due to the presence of WMHs in the majority of the patients in these populations. This pattern was also visible to a lesser extent as hypointensity in the T1w templates, as well as hyperintensity in the T2w, PD, and T2* templates (Figures 3 to 5).

**Figure 1.**
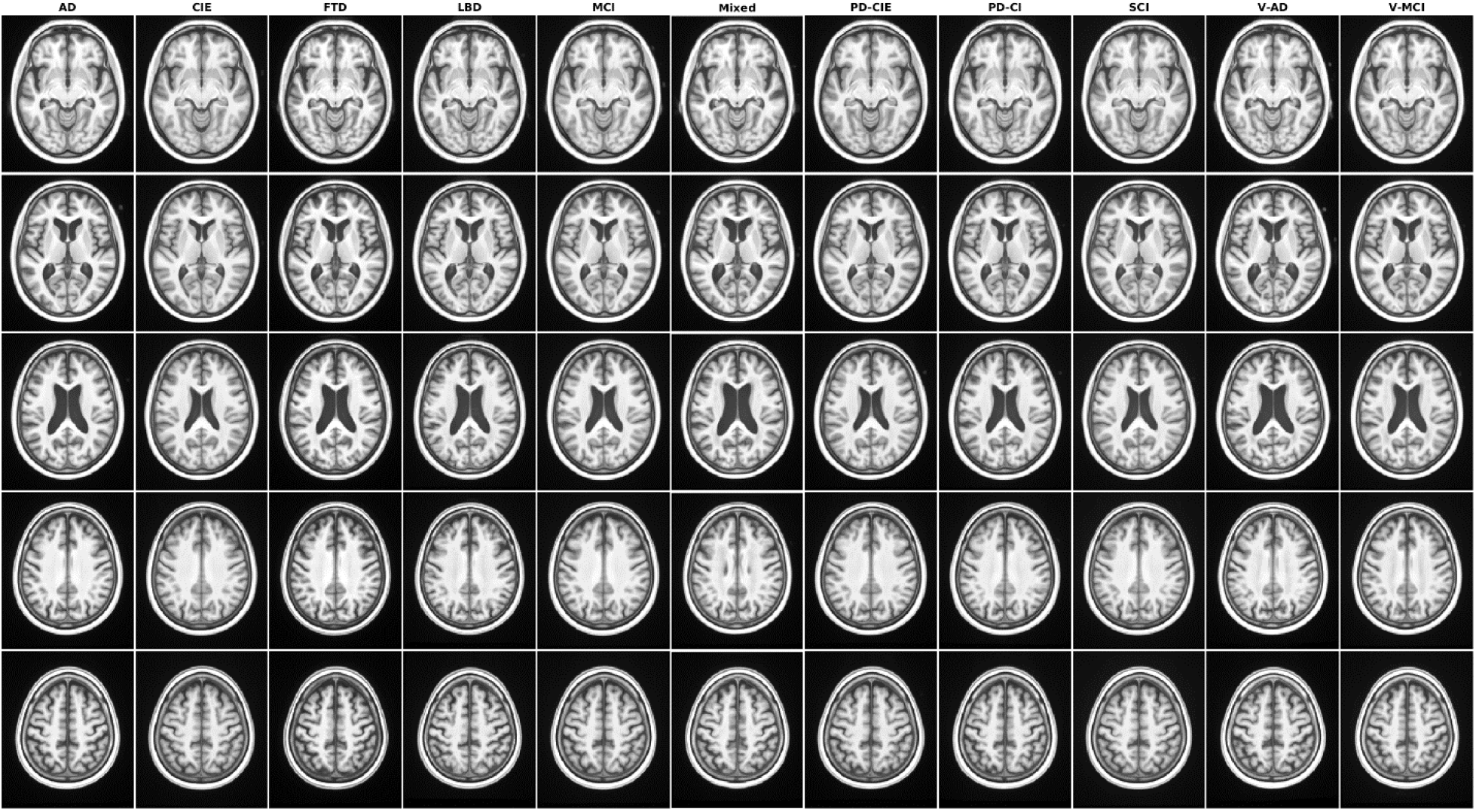
Axial slices of T1w average templates for all diagnostic groups.

**Figure 2.**
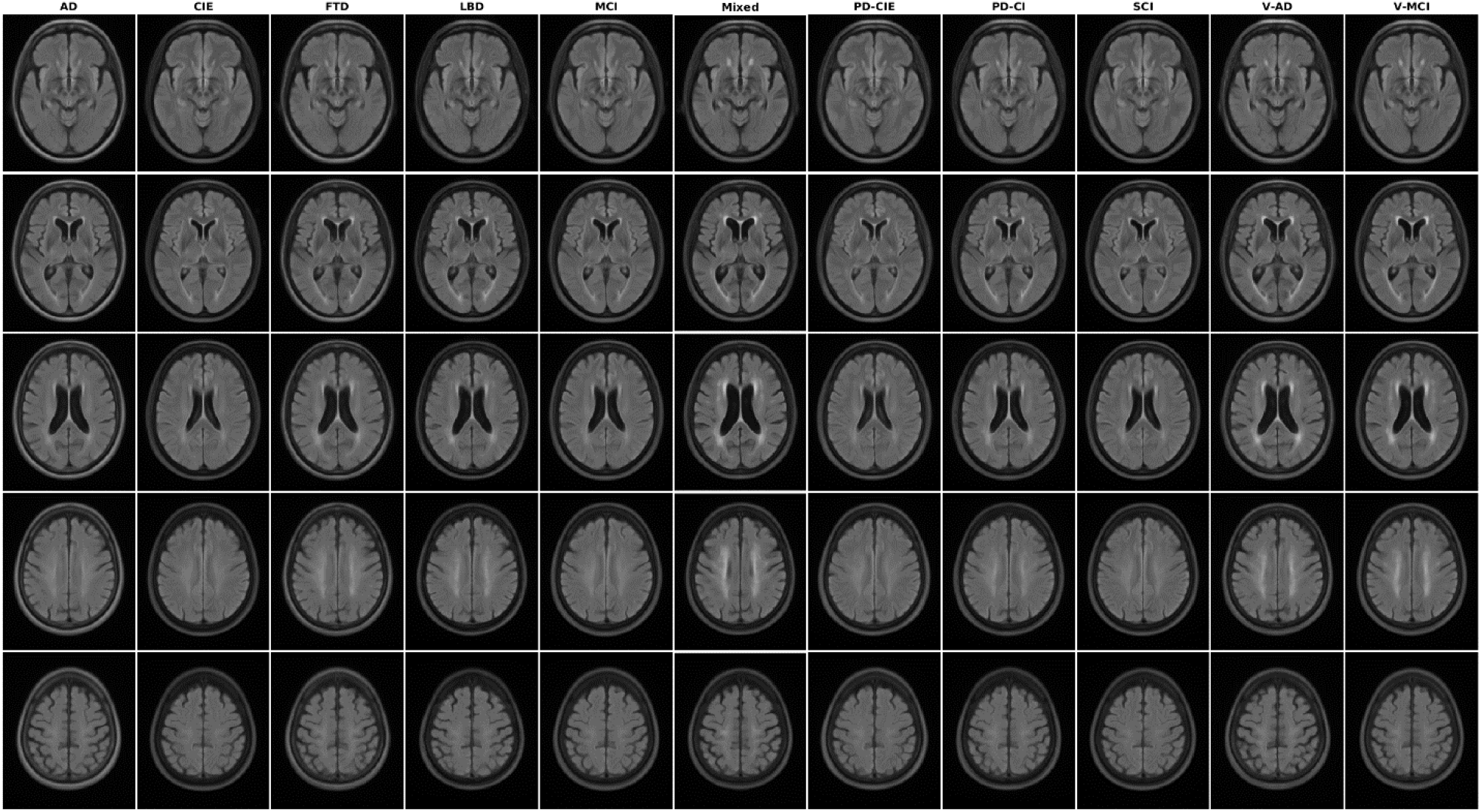
Axial slices of FLAIR average templates for all diagnostic groups.

**Figure 3.**
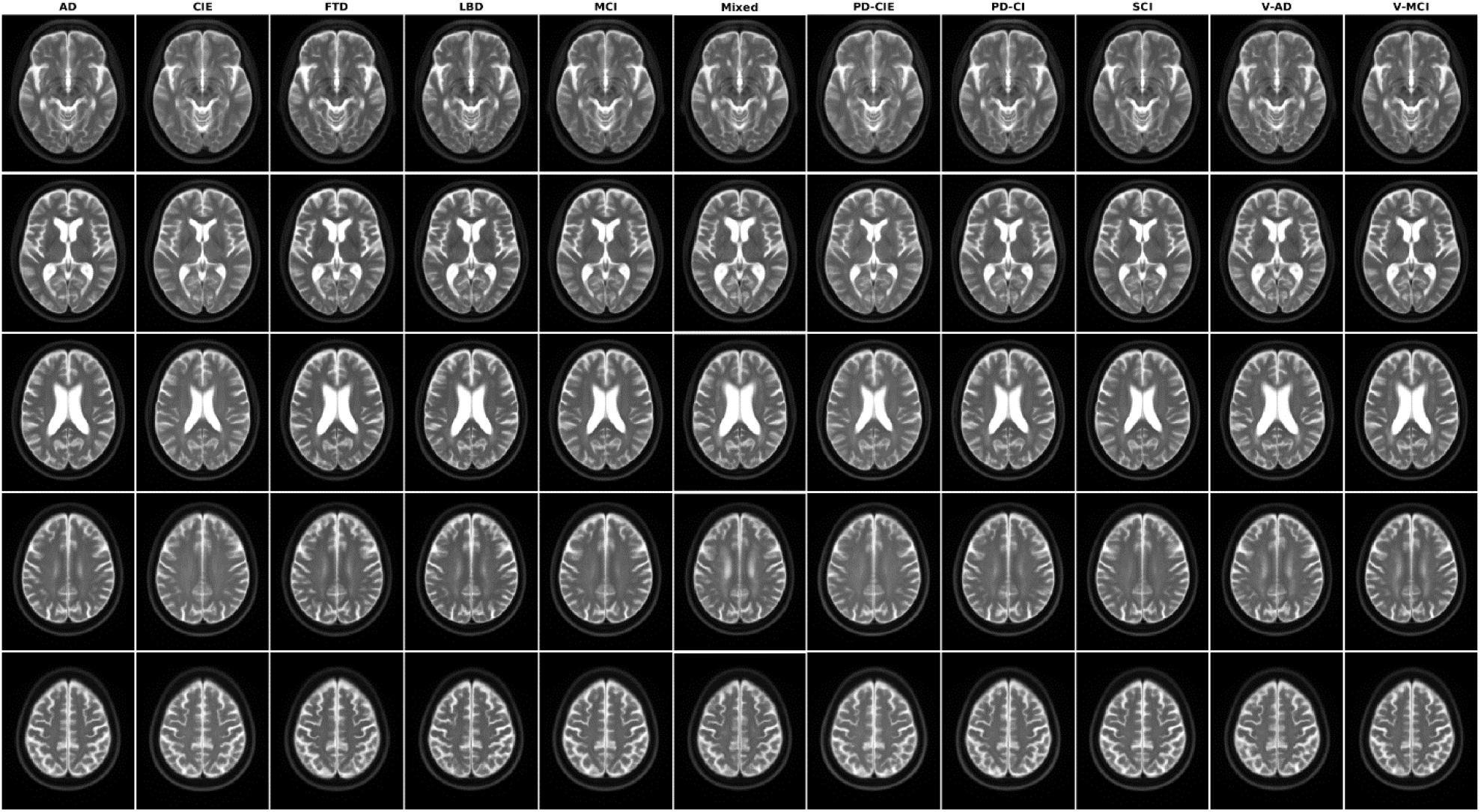
Axial slices of T2w average templates for all diagnostic groups.

**Figure 4.**
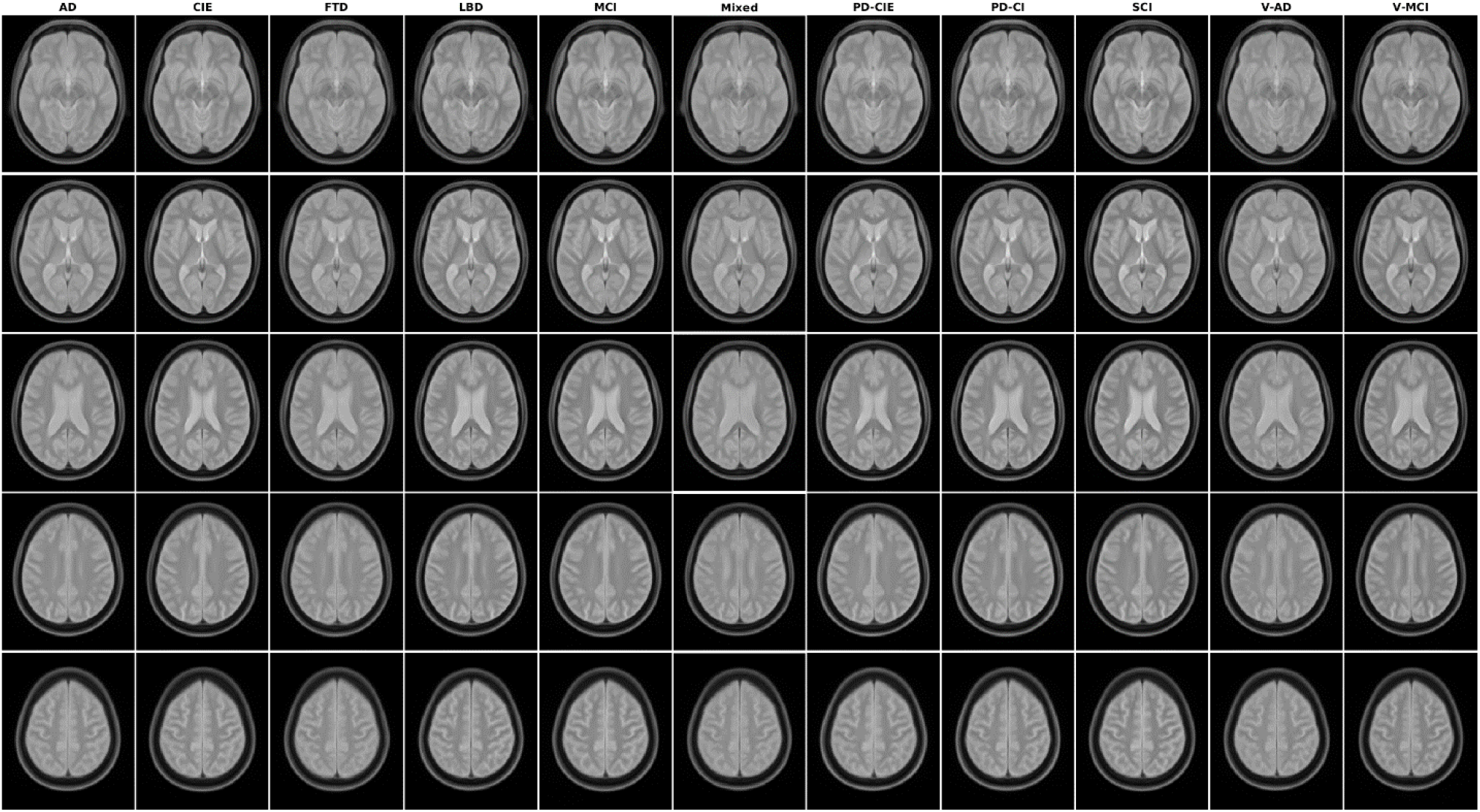
Axial slices of PD average templates for all diagnostic groups.

**Figure 5.**
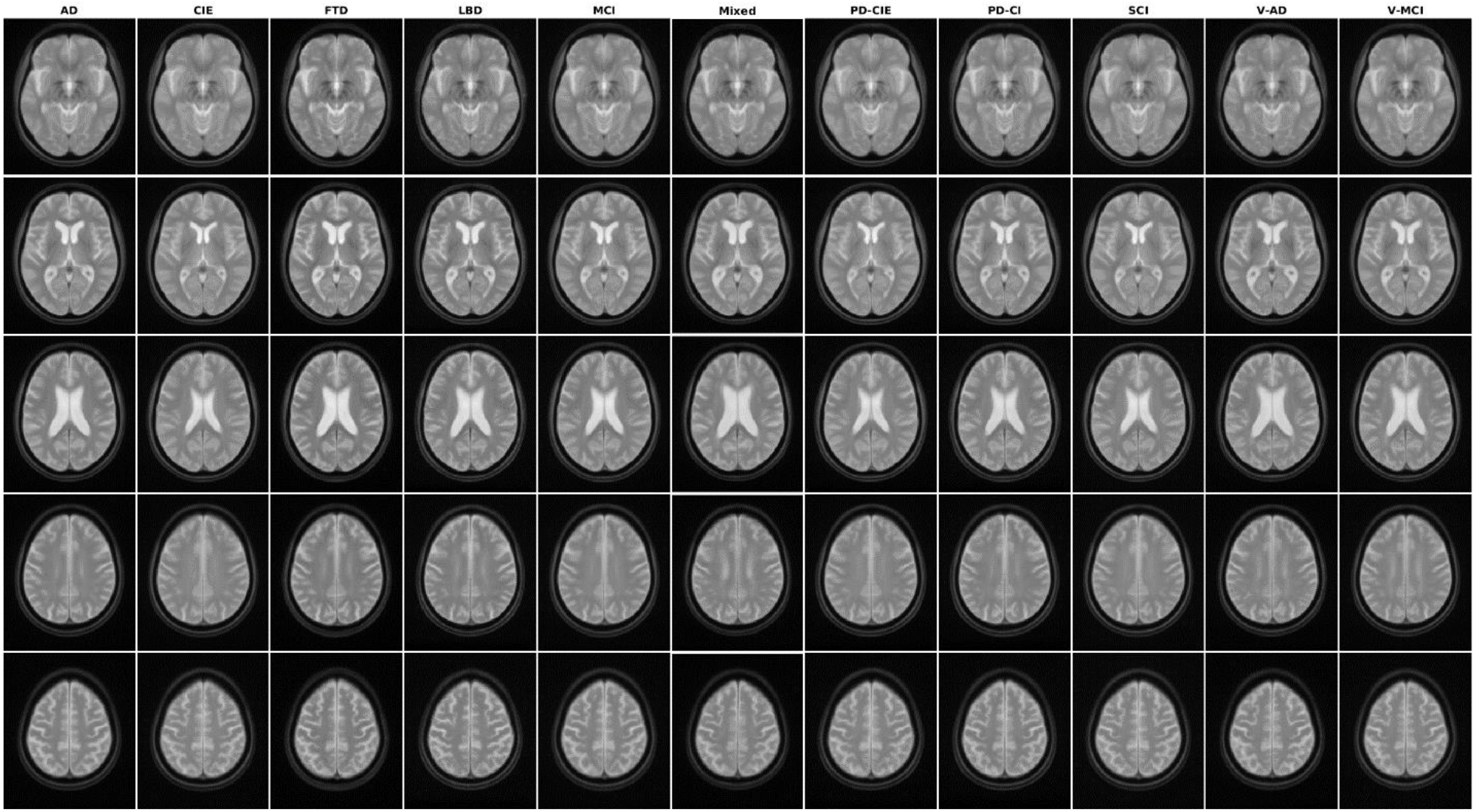
Axial slices of T2* average templates for all diagnostic groups.

Figure 6 shows axial slices of the male and female templates for all diagnostic groups and sequences. Overall, male templates have larger ventricles and greater levels of atrophy than female templates. For more detailed figures of each template, see the supplementary materials (Figures S.12-S.31).

**Figure 6.**
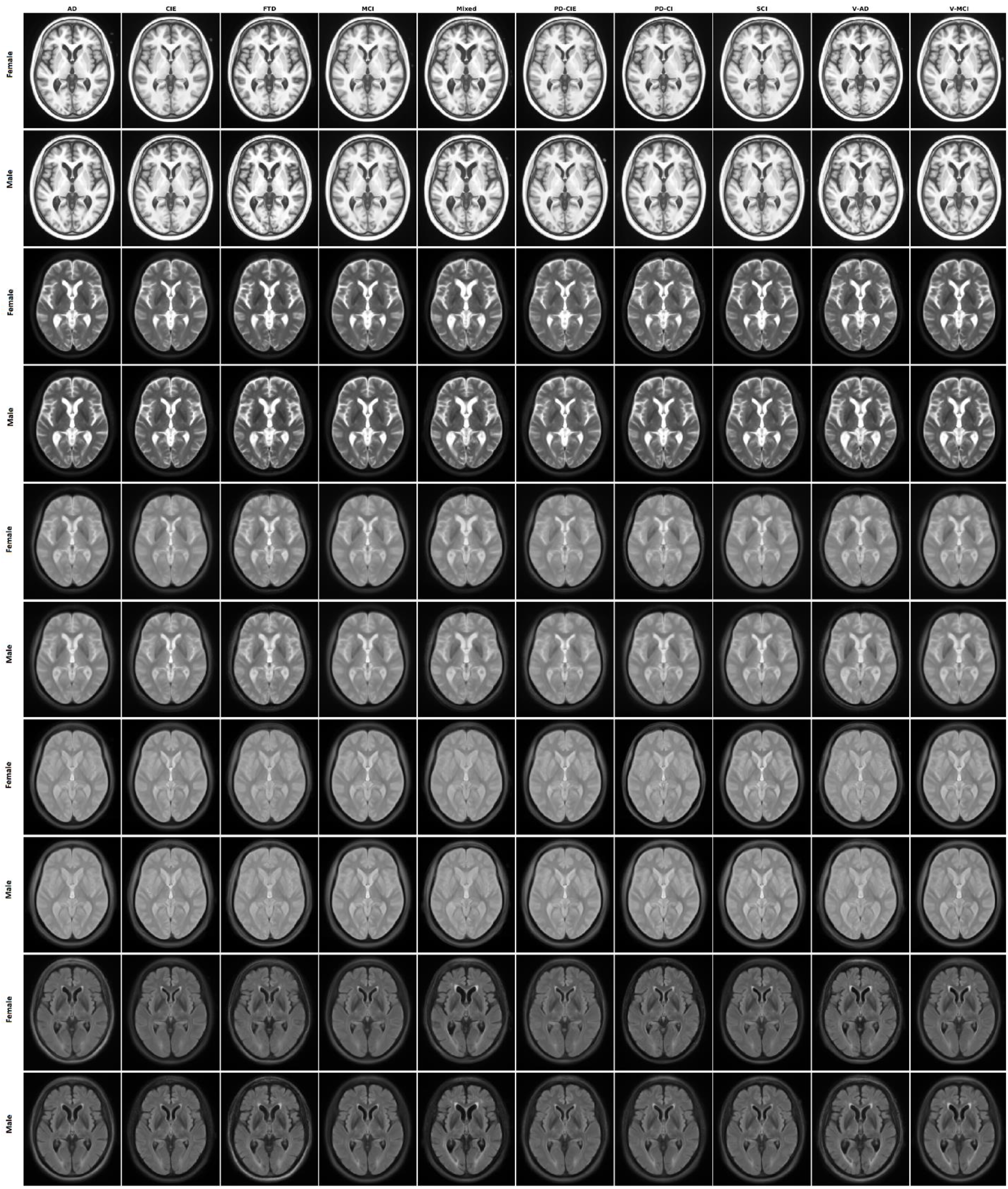
Axial slices of average male and female templates for all sequences and diagnostic groups.

Figure 7 shows axial slices of the templates for the human phantom (SIMON).

**Figure 7.**
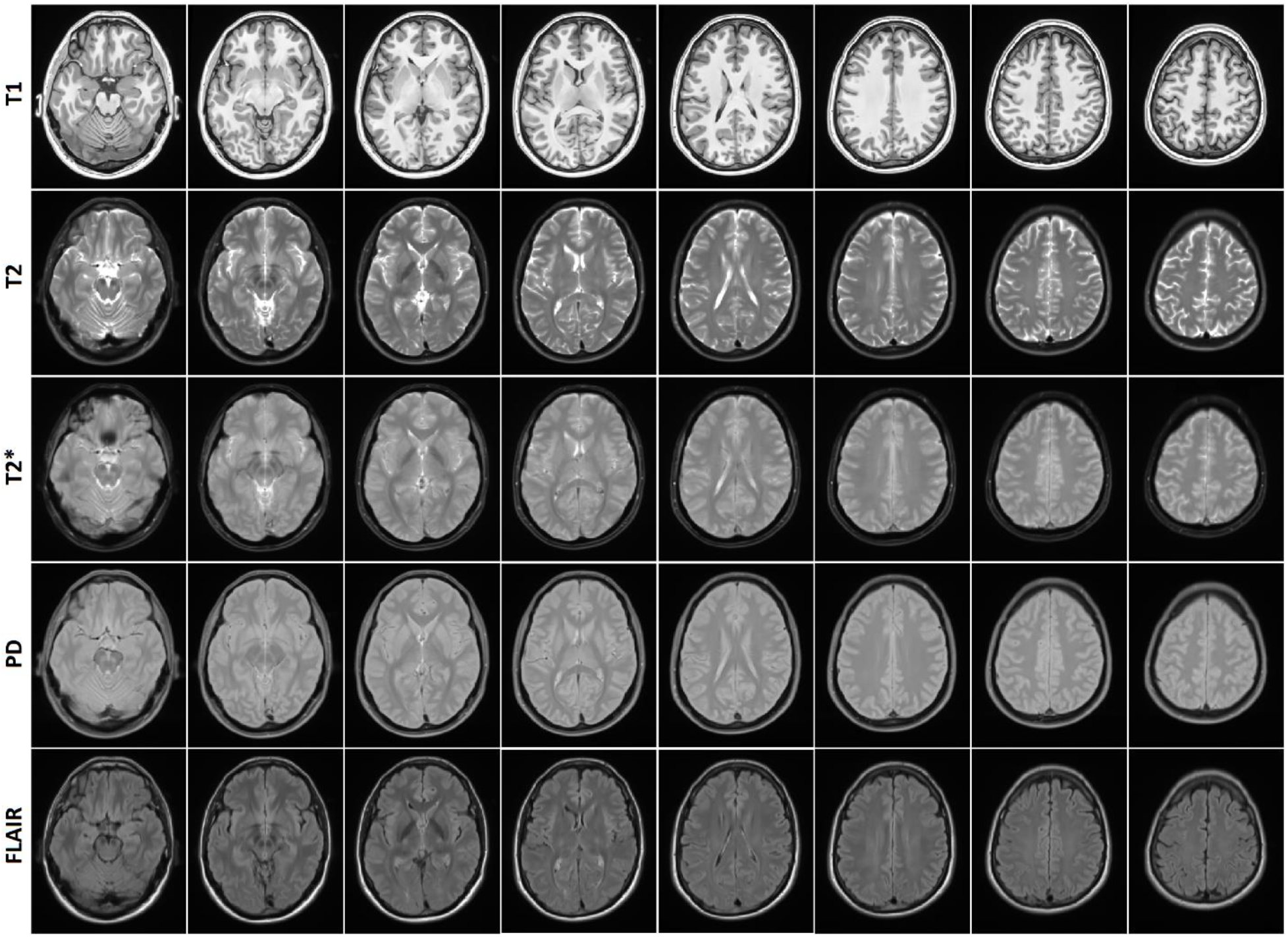
Axial slices of human phantom (SIMON) templates for all sequences.

#### Volumetric Comparisons

Tables 2, 3, and 4 summarize the grey and white matter (GM, WM) and cerebrospinal fluid (CSF) volumetric information for the templates as segmented by FreeSurfer. Figure 8 compares GM volumes (log transformed) of each template against the CIE template. Data points below the reference line (shown in red) indicate lower values for the template in comparison with the CIE template. As expected, cognitively impaired and dementia templates had lower GM values than the CIE template, whereas both cognitively intact PD-CIE and SCI templates had similar volumes to the CIE template (i.e. data points fall on the reference line).

**Table 3.**
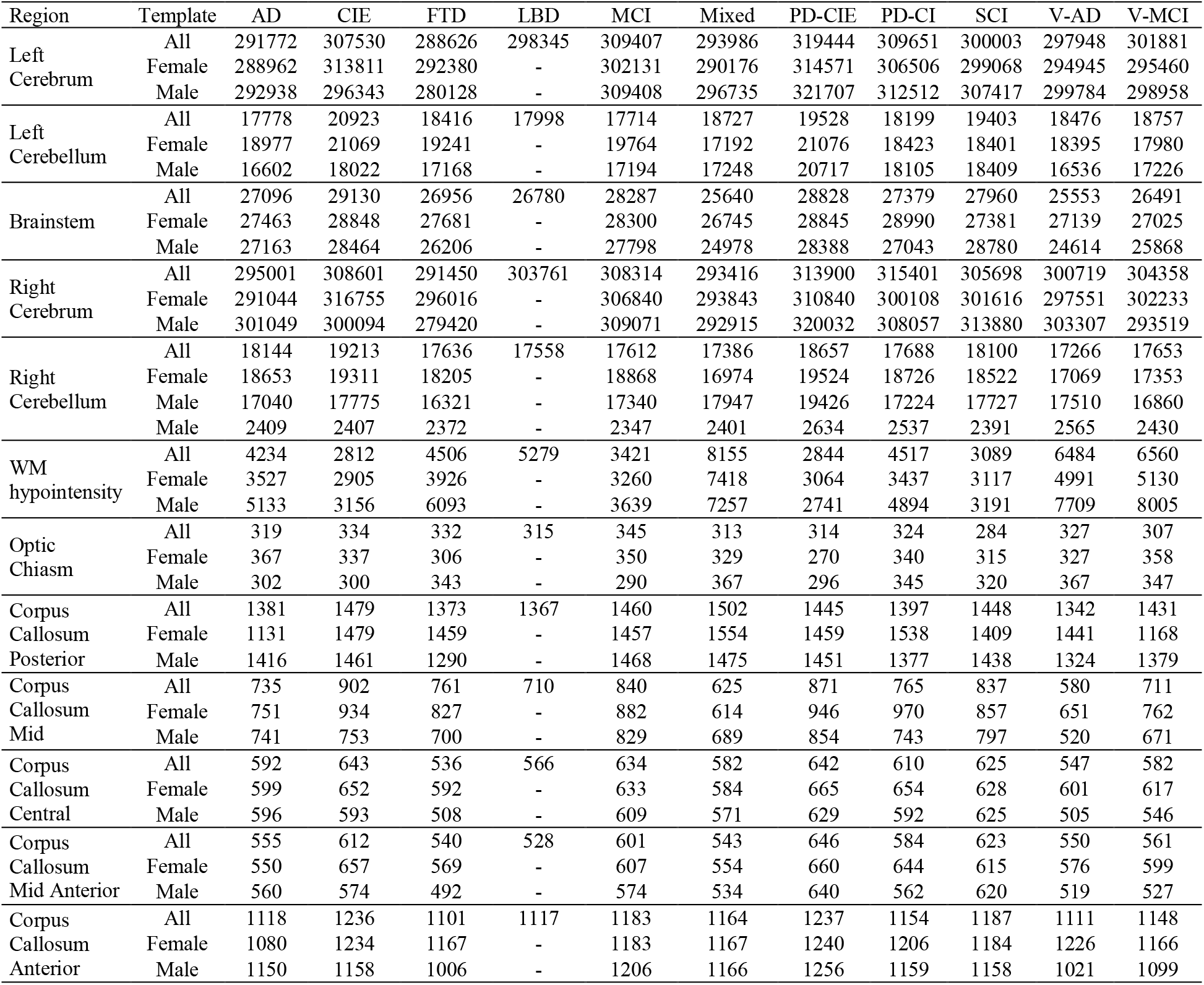
Volumetric WM information (in mm^3^) for each template based on FreeSurfer segmentations.

**Table 4.**
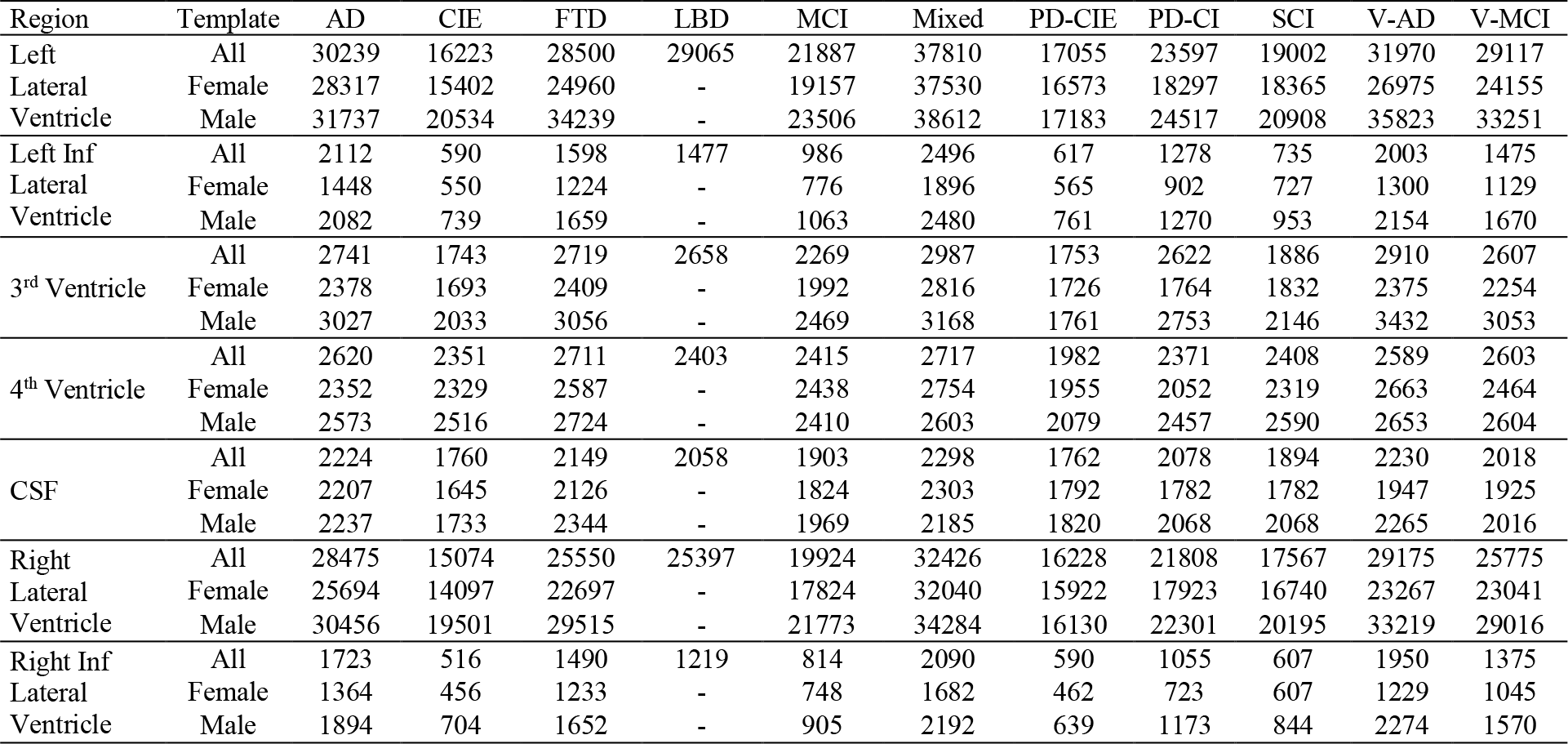
Volumetric CSF information (in mm^3^) for each template based on FreeSurfer segmentations.

**Figure 8.**
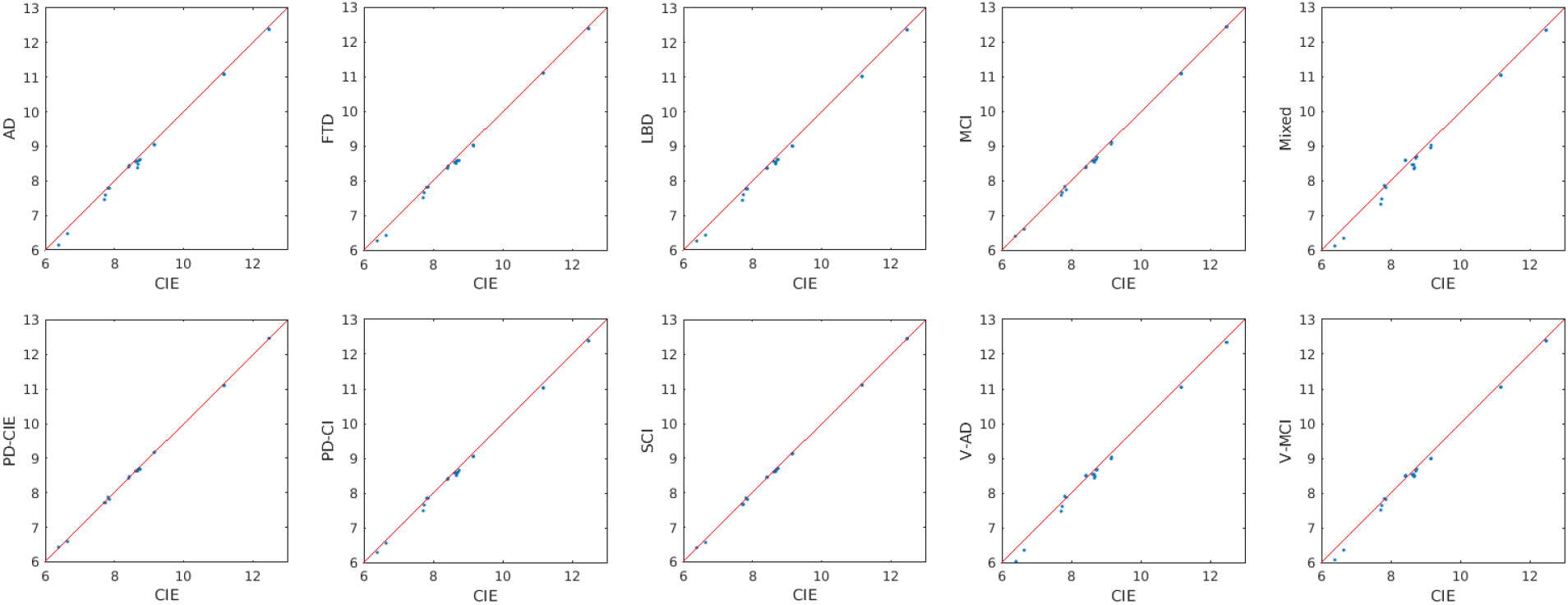
FreeSurfer based GM volumes for each diagnostic group versus the CIE template. CIE= Cognitively Intact Elderly.

Figure 9 compares GM volumes (log transformed) of male versus female templates. Note that since all templates have been linearly registered to the MNI-ICBM2009c template prior to the template creation step, all volumetric values reflect variabilities after accounting for intracranial volume differences and are not caused by potential head size differences between males and females. Data points below the reference line (shown in red) indicate lower values for the male template in comparison with the female template. In the AD and mixed templates, the nucleus accumbens areas bilaterally had lower volumes in the male templates. In the PD-CI template, most regions had slightly lower GM volumes in the male template.

**Figure 9.**
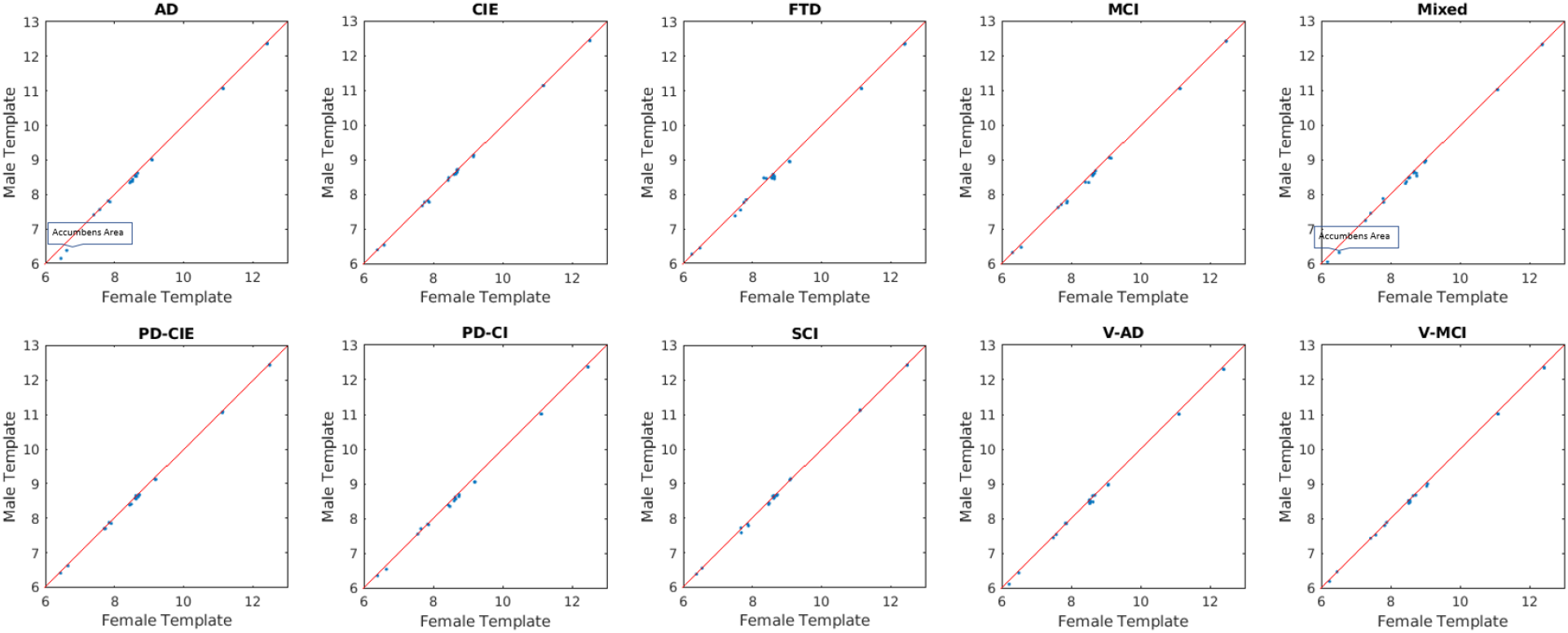
FreeSurfer based GM volumes for male and female templates for each diagnostic group.

As expected, mixed dementia, vascular MCI, and vascular AD templates had higher WM hypointensity volumes (corresponding to the WMHs on FLAIR and T2w sequences) on T1w templates (Table 3). Male templates for AD, FTD, PD-CI, V-MCI, and V-AD also had greater WM hypointensity volumes than the female templates (Table 3). The mixed template had the largest ventricles (Table 4), followed by V-AD and AD templates. As expected, CIE template had the smallest ventricles, followed by PD-CIE, and SCI. In all diagnostic groups, lateral ventricles were larger for the male templates in comparison with the female templates. This difference was most prominent in the V-AD group, for which the left and right lateral ventricles were 33% and 43% larger respectively for the male template (Table 4). Regarding asymmetry, in the FTD, V-MCI, and mixed templates, the left lateral ventricle was 12%, 13%, and 17% larger than the right lateral ventricle. This difference was more prominent in the male templates for FTD and V-MCI groups, whereas for the mixed group, the female template had greater asymmetry in the ventricles. All of these differences highlight the need for group-specific templates in multi-individual, multi-centric studies.

**Table 2.**
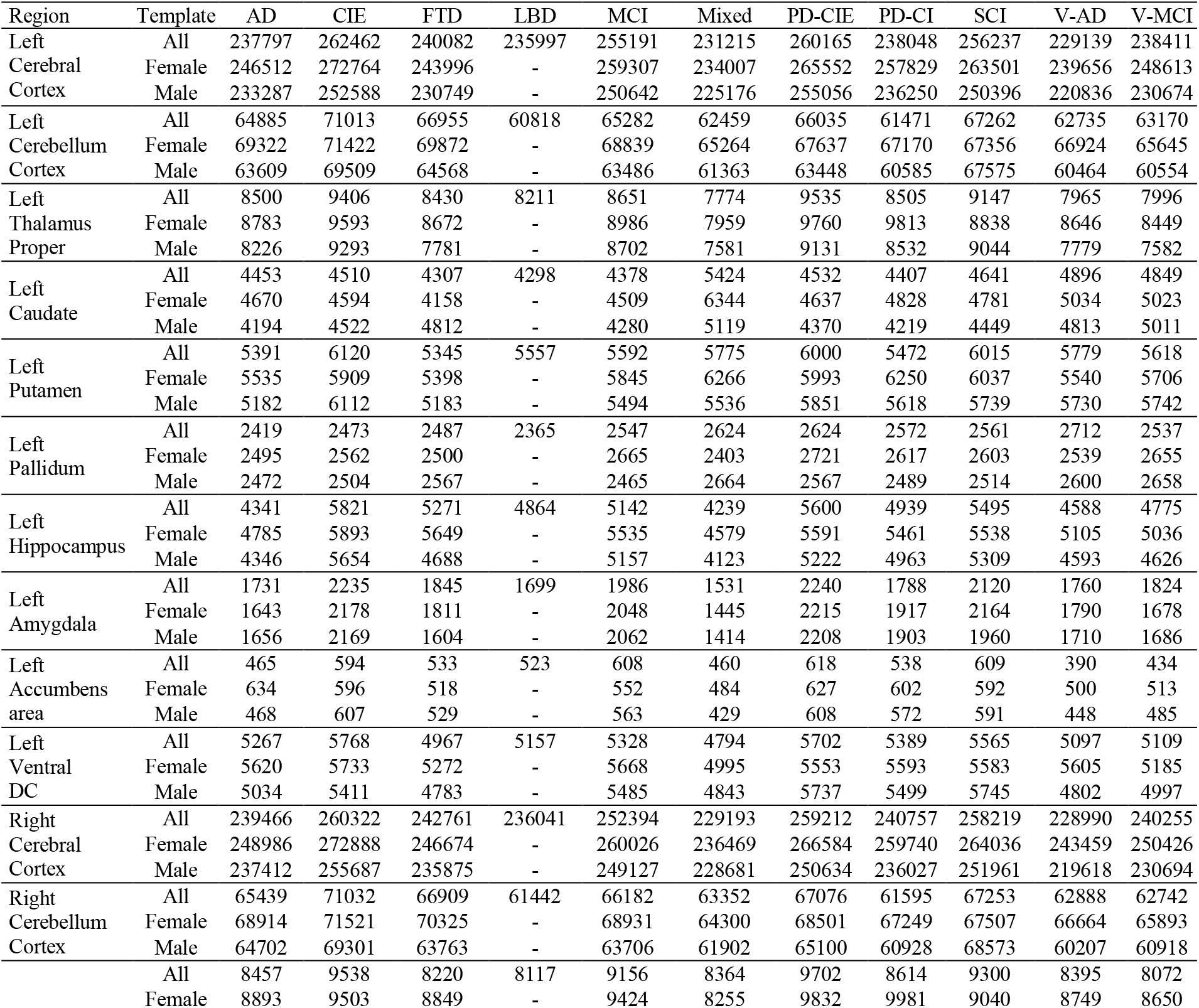
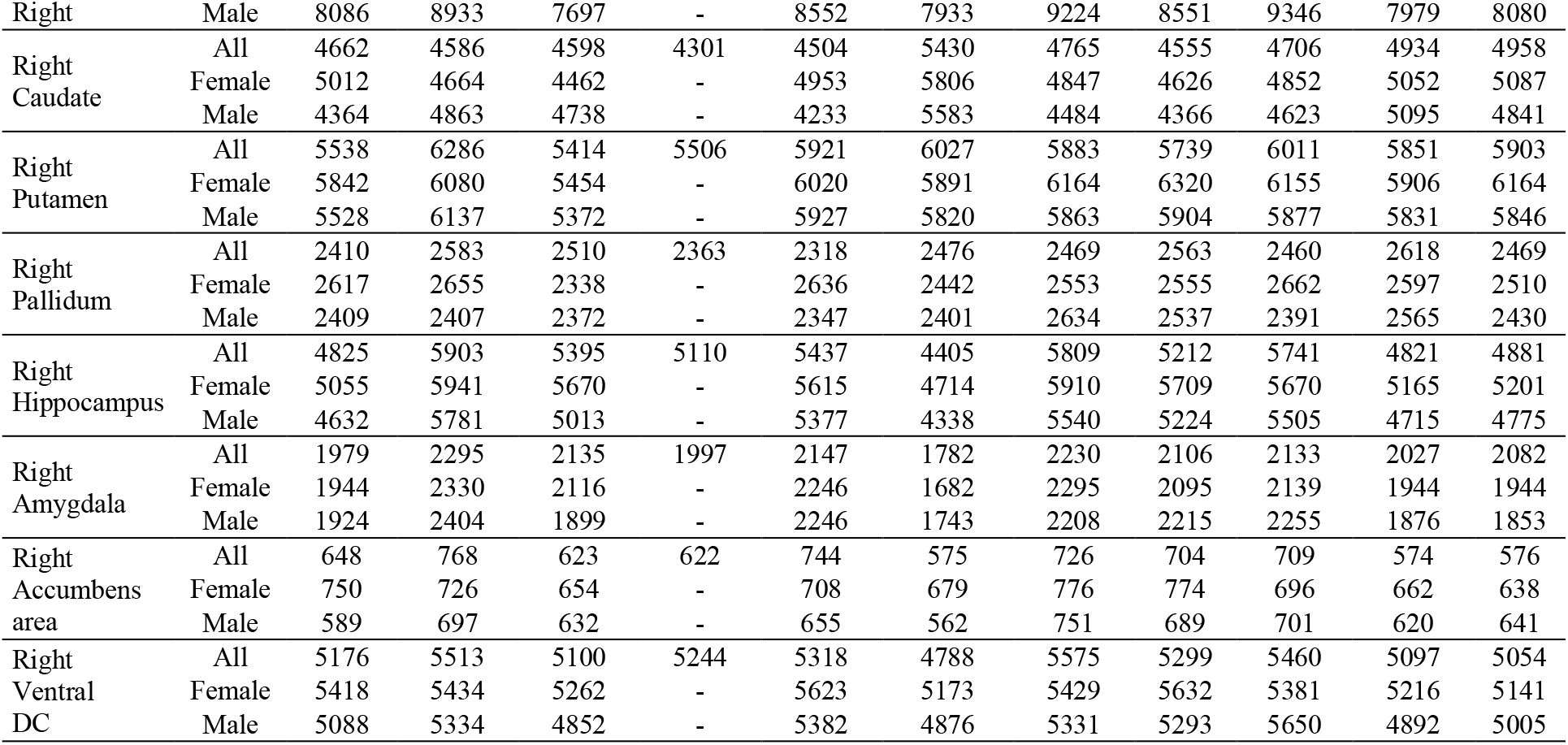
Volumetric GM information (in mm^3^) for each template based on FreeSurfer segmentations.

## Data Availability

The average template files for all groups and sequences are available in both compressed MINC and NIfTI formats at G-Node (https://gin.g-node.org/mahsadadar/CDIP_Templates) as well as Zenodo (https://zenodo.org/record/5018356#.YNPNkExE3b0).

## Code Availability

The scripts for generating unbiased average templates are publicly available at https://github.com/vfonov/nist_mni_pipelines.

## Authors Contributions

**Mahsa Dadar**: Study concept and design, analysis of the data, drafting and revision of the manuscript.

**Richard Camicioli:** Study concept and design, interpretation of the data, revising the manuscript.

**Simon Duchesne:** Study concept and design, interpretation of the data, revising the manuscript.

## Acknowledgements

MD is supported by a scholarship from the Canadian Consortium on Neurodegeneration in Aging (CCNA) as well as by an Alzheimer Society Research Program (ASRP) postdoctoral award. CCNA is supported by a grant from the Canadian Institutes of Health Research with funding from several partners.

## Competing Interests

The authors declare no competing interests.

### Abbreviations

AD: Alzheimer’s Dementia
CIE: Cognitively Intact Elderly
FLAIR: FLuid Attenuated Inversion Recovery
FTD: Fronto-temporal Dementia
LBD: Lewy Body Dementia
MCI: Mild Cognitive Impairment
MRI: Magnetic Resonance Imaging
PD: Parkinson’s Disease
PD: Proton Density
SCI: Subjective Cognitive Impairment
T1w: T1-weighted
T2w: T2-weighted
TE: echo time
TI: inversion time
TR: repetition time
V-AD: Vascular Alzheimer’s Dementia
V-MCI: Vascular Mild Cognitive Impairment
WMH: White Matter Hyperintensity

## Notes

### Competing Interest Statement

The authors have declared no competing interest.

### Author Declarations

The COMPASS-ND study was approved by the Jewish General Hospital Research Ethics Board. Written informed consent was obtained from all participants.

